# A Comparison of the Randomized Clinical Trial Efficacy and Real-World Effectiveness of Tofacitinib for the Treatment of Inflammatory Bowel Disease: A Cohort Study

**DOI:** 10.1101/19007195

**Authors:** Vivek A. Rudrapatna, Benjamin S. Glicksberg, Atul J. Butte

## Abstract

**Background:** Real-world data are receiving attention from regulators, biopharmaceuticals and payors as a potential source of clinical evidence. However, the suitability of these data to produce evidence commensurate with randomized controlled trials (RCTs) and the best practices in their use remain unclear. We sought to compare the real-world effectiveness of Tofacitinib in the treatment of IBD against efficacy rates published by corresponding RCTs.

**Methods:** Electronic health records at the University of California, San Francisco (UCSF) were queried and reviewed to identify 86 Tofacitinib-treated IBD patients through 4/2019. The primary endpoint was treatment effectiveness. This was measured by time-to-treatment-discontinuation and by the primary endpoints of RCTs in Ulcerative Colitis (UC) and Crohn’s Disease (CD). Endpoints were measured and analyzed following a previously published protocol and analysis plan.

**Findings:** 86 patients (68 with UC, 18 with CD) initiated Tofacitinib for IBD treatment. Most of the data needed to calculate baseline and follow-up disease activity indices were documented within the EHR(77% for UC, 91% for CD). Baseline characteristics of the UCSF and RCT cohorts were similar, except for a longer disease duration and 100% treatment failure of Tumor Necrosis Factor inhibitors in the former. None of the UCSF cohort would have met the RCT eligibility criteria due to multiple reasons.

The rate of achieving the RCT primary endpoints were highly similar to the published rates for both UC(16%, P=0·5) and CD (38%, P=0·8). However, treatment persistence was substantially higher: 69% for UC (week 52) and 75% for CD (week 26).

**Interpretation:** An analysis of routinely collected clinical data can reproduce published Tofacitinib efficacy rates, but also indicates far greater treatment durability than suggested by RCTs including possible benefit in CD. These results underscore the value of real-world studies to complement RCTs.

**Funding:** The National Institutes of Health and UCSF Bakar Institute

**Research in Context:** *Evidence before this study:* Tofacitinib is the most recently approved treatment for Ulcerative Colitis. Data related to treatment efficacy for either IBD subtype is generally limited, whether from controlled trials or real-world studies. A search of clinicaltrials.gov was performed in January 2019 for completed phase 2 or 3, interventional, placebo-controlled clinical trials matching the terms “Crohn’s Disease” OR “Ulcerative Colitis” in the conditions field, and matching “Placebo” AND “Tofacitinib” OR “CP-690,550”) in the Interventions field. We identified three Phase 3 trials for UC (OCTAVE trials, all initially reported in a single article in 2016) and three Phase 2 trials of CD (two published in the same article in 2017, one reported in 2014). The Phase 3 UC trials reported 57·6% pooled clinical response rate in the Tofacitinib-assigned groups after 8 weeks (induction), and a 37·5% pooled remission rate among eligible induction trial responders in the Tofacitinib-assigned groups at 52 weeks. The 2017 CD trial reported a 70·8% pooled rate of response or remission in the Tofacitinib-assigned groups after 8 weeks, and a 47·6% pooled rate of response or remission among enrolled induction-trial responders at 26 weeks. A bias assessment of both UC and CD trials indicated a high risk of attrition bias and unclear risk of bias related to conflicts of interest. We also performed a search of pubmed.gov in January 2019 using search terms (“Colitis” OR “Crohn’s”) AND (“Tofacitinib” OR “CP-690,550”) OR “real-world” to identify cohort studies of Tofacitinib efficacy in routine clinical practice. No studies meeting these criteria were identified.

*Added value of this study:* This is one of the early studies to closely compare the results of clinical trials with the continuously-updated data captured in the electronic health records, and the very the first to assess the efficacy-effectiveness gap for Tofacitinib. We found that none of the patients treated at our center thus far would have qualified for the clinical trial based on published eligibility criteria. We found that the drug appeared to perform similarly to its efficacy when using the endpoints reported in clinical trials, but treatment persistence was significantly greater than would have been expected from the reported trial outcomes: 69% for UC at week 52 and 75% for CD at week 26.

*Implications of all the available evidence:* Tofacitinib is an effective treatment for the Ulcerative colitis and may be efficacious for Crohn’s disease. Controlled trials may not be representative of real-world cohorts, may not be optimally designed to identify efficacious drugs, and may not accurately predict patterns of use in clinical practice. Further studies using real-world data as well as methods to enable their proper use are needed to confirm and continuously monitor the efficacy and safety of drugs, both for on- and off-label use.

## Introduction

Inflammatory Bowel Disease (IBD) has increasingly been recognized as a global disease with accelerating incidence and prevalence in newly industrialized nations.^1^ Although IBD has historically been quite morbid – associated with progressive bowel damage, malnutrition, chronic pain and neoplasia – recent decades have seen a revolution in disease treatment and natural history with the advent of molecularly targeted therapies.

Most of these new treatments have been monoclonal antibody biologics: agents that are expensive to repare and deliver, and associated with a loss of response over time. The first oral small-molecule for the treatment of moderate to severe Ulcerative Colitis (UC) was approved by the EMA and the US FDA in 2018. Tofacitinib was approved on the basis of positive Phase 3 studies where it showed a statistically significant reduction in the Mayo score over placebo by week 52.^2^ Tofacitinib was also studied in two Phase IIb randomized controlled trials (RCTs) of Crohn’s Disease (CD), the other major subtype of IBD. In CD however, no statistical difference from placebo was seen in Crohn’s Disease Activity Index (CDAI) reduction at week 26.^3^ Consequently, further investigation of Tofacitinib in Crohn’s Disease was abandoned.

RCTs have long been considered the gold standard for clinical evidence.^4^ However, these studies have come under greater scrutiny due to questions of their generalizability to the average clinical setting. In particular, they have been associated with restrictive eligibility criteria that would exclude many real-world patients being considered for care.^5^ Moreover, RCTs often do not measure the very same endpoints as those used in routine clinical care. In practice the decision to continue, modify, or discontinue treatment typically requires a patient-centric and setting-specific discussion of risks, benefits, and alternatives. As such, RCT endpoints which are commonly binary, uniformly applied, and analyzed according to the intention-to-treat principle are often less apt at answering common patient questions such as: “What are the chances that this treatment is going to work?”

Recent years have seen significant interest in *real-world data* (RWD) in part due to their potential to answer these sorts of pragmatic questions.^6–8^ However, their use has been fraught with many challenges including missing data and misclassification. Widely-adopted standards for the handling of this data have not been clearly established, nor is careful benchmarking against other gold-standard sources of evidence presently a common practice.

In this cohort study we attempt to answer the following questions: Is routinely collected clinical data complete enough to measure the endpoints assessed in IBD RCTs of Tofacitinib? Using both these regulatory endpoints as well as the more pragmatic measure of *time to treatment discontinuation*, what is the real-world effectiveness of Tofacitinib? How does it compare to trial efficacy rates? Are there any systematic differences between the populations under study in both controlled and real-world settings?

## Methods

This study was performed in accordance with the STROBE statement^9^ (See Supplemental Content).

### Patient identification and covariate extraction

We queried a structured database of deidentified Electronic Health Records (EHR) at the University of California, San Francisco (UCSF) to identify patients meeting the following criteria: 1) age 18 or older, 2) presence of a medication order for Tofacitinib, and 3) presence of an IBD diagnosis code (ICD-10-CM K50*/K51*) assigned during a Gastroenterology clinic visit (Table 1). The scope of the search extended from the instantiation of the EHR software (6/2012) through the time of the query (4/2019). We obtained Institutional Review Board approval (#18-24588) to obtain identifiable patient record data, to confirm that these patients had initiated treatment on Tofacitinib for the treatment of IBD, and to assess treatment compliance. Compliance was defined as adherence to the treatment plan (e.g. dose, frequency, duration) determined by the ordering provider.

**Table 1:** Demographic comparison of subjects studied in RCTs of Ulcerative Colitis and Crohn’s Disease with subjects assigned to receive Tofacitinib and corresponding patients at UCSF. *: *Includes patients who received intravenous corticosteroids at the time of Tofacitinib initiation*.

|  | OCTAVE Induction 1<br>(n=475) | Sample of UC Cohort<br>(n=28) |  | Panés et al. 2017<br>(n=86) | Sample of CD Cohort<br>(n=13) |
| --- | --- | --- | --- | --- | --- |
| **Male sex - no. (%)** | 277 (58.2) | 16 (57) | **Female, n (%)** | 47 (54.7) | 9 (69.2) |
| **Age – yr** | 41.3 ± 14.1 | 43.2 ± 14.4 | **Age, years** |  |  |
| **Duration of disease - yr** |  |  | -- Mean (SD) | 39.3 (13.7) | 39.7 (19.5) |
| -- Median | 6.5 | 10.2 | **Weight, kg** |  |  |
| -- Range | 0.3-42.5 | 2.2-51.4 | -- Mean (SD) | 71.6 (18.8) | 69.9 (16.3) |
| **Extent of Disease -**<br>**no./total no. (%)** |  |  | **Race, n (%)** |  |  |
| -- Proctosigmoiditis | 64/475 (13.7) | 3/28 (10.7) | -- White | 72 (83.7) | 9 (69.2) |
| -- Left-sided colitis | 158/475 (33.3) | 6/28 (21.4) | -- Black | 2 (2.3) | 0 (0) |
| -- Extensive colitis or<br>pancolitis | 252/475 (53.1) | 19/28 (67.9) | -- Asian | 11 (12.8) | 1 (7.7) |
| **Total Mayo score** | 9.0 ± 1.4 | 8.5 ± 1.8 | -- Others | 1 (1.2) | 0 (0) |
| **Partial Mayo score** | 6.3 ± 1.2 | 6 ± 1.6 | **Duration since CD diagnosis, years** |  |  |
| **C-reactive protein -**<br>**mg/liter** |  |  | -- Mean (SD) | 11.3 (9.7) | 14.4 (8.2) |
| -- Median | 4.4 | 5.8 | **Extent of disease, n (%)** |  |  |
| -- Range | 0.1-208.4 | 0.8-70.6 | -- L1 (I/TI) | 7 (8.1) | 1 (7.7) |
| **Glucocorticoid use at**<br>**baseline\*** | 214 (45.0) | 17 (60.7) | -- L1/4 (I/TI + UGI) | 2 (2.3) | 2 (15.4) |
| **Previous treatment with**<br>**TNF antagonist – no. (%)** | 254 (53.4) | 28 (100) | -- L2 (C) | 5 (5.8) | 0 (0) |
| **Previous treatment failure -**<br>**no. (%)** |  |  | -- L2/4 (C + UGI) | 16 (18.6) | 1 (7.7) |
| -- TNF antagonist | 243 (51.1) | 28 (100) | -- L3 (IC) | 15 (17.4) | 4 (30.8) |
| -- Glucocorticoid | 350 (73.5) | 24 (85.7) | -- L3/4 (IC + UGI) | 39 (45.3) | 5 (38.5) |
| -- Immunosuppressant | 360 (75.6) | 21 (75) | **Prior use of TNFi, n (%)** | 66 (76.7) | 13 (100) |
|  |  |  | **Use of corticosteroids at study**<br>**entry, n (%)** | 28 (32.6) | 7 (53.8) |
|  |  |  | **Baseline CDAI score** |  |  |
|  |  |  | -- Mean (SD) | 320 (61.66) | 374 (183.73) |
|  |  |  | **Baseline CRP, mg/L** |  |  |
|  |  |  | -- Median (min-max) | 5.5 (0.2-126) | 28.7 (3.5-107) |

Following a openly published protocol and statistical analysis plan,^10^ we reviewed all patient records to measure the time to treatment discontinuation or last known use. We selected a subset of these records to perform more detailed covariate extraction from the EHR following the aforementioned protocol. These covariates included baseline demographics as well as the primary endpoints of the Phase 2b/3 RCTs of Tofacitinib for CD^3^ and UC^2^ respectively. The time windows used for covariate extraction were months -6 through 0 for the baseline data, and months 2 through 8 for the follow-up data. We selected these windows in order to balance data availability and typical practice patterns with comparability to protocol-driven endpoint measurements in RCTs.

### Data quality and missing data assessments

We assessed the quality of the data in detail prior to proceeding with further analysis. We confirmed the basic epidemiological properties of the dataset, such as the bimodal age of IBD onset which has been previously reported^11,12^ (Supplemental Content eFigure 1). We annotated missing data and characterized its distribution. Given that missing data was present across several variables following different non-normal distributions, we performed multiple imputation by chained equations using random forest classification/regression models (20 Markov chains, 10 iterations). We augmented our imputation model with endpoints of interest in order to facilitate ‘congeniality’.^13^ We used all available variables as auxiliary variables to satisfy the missing at random (MAR) assumption and increase imputation power. We examined trace plots and strip plots to confirm Markov chain convergence and the plausibility of imputed values respectively (Supplement).

**Figure 1:**
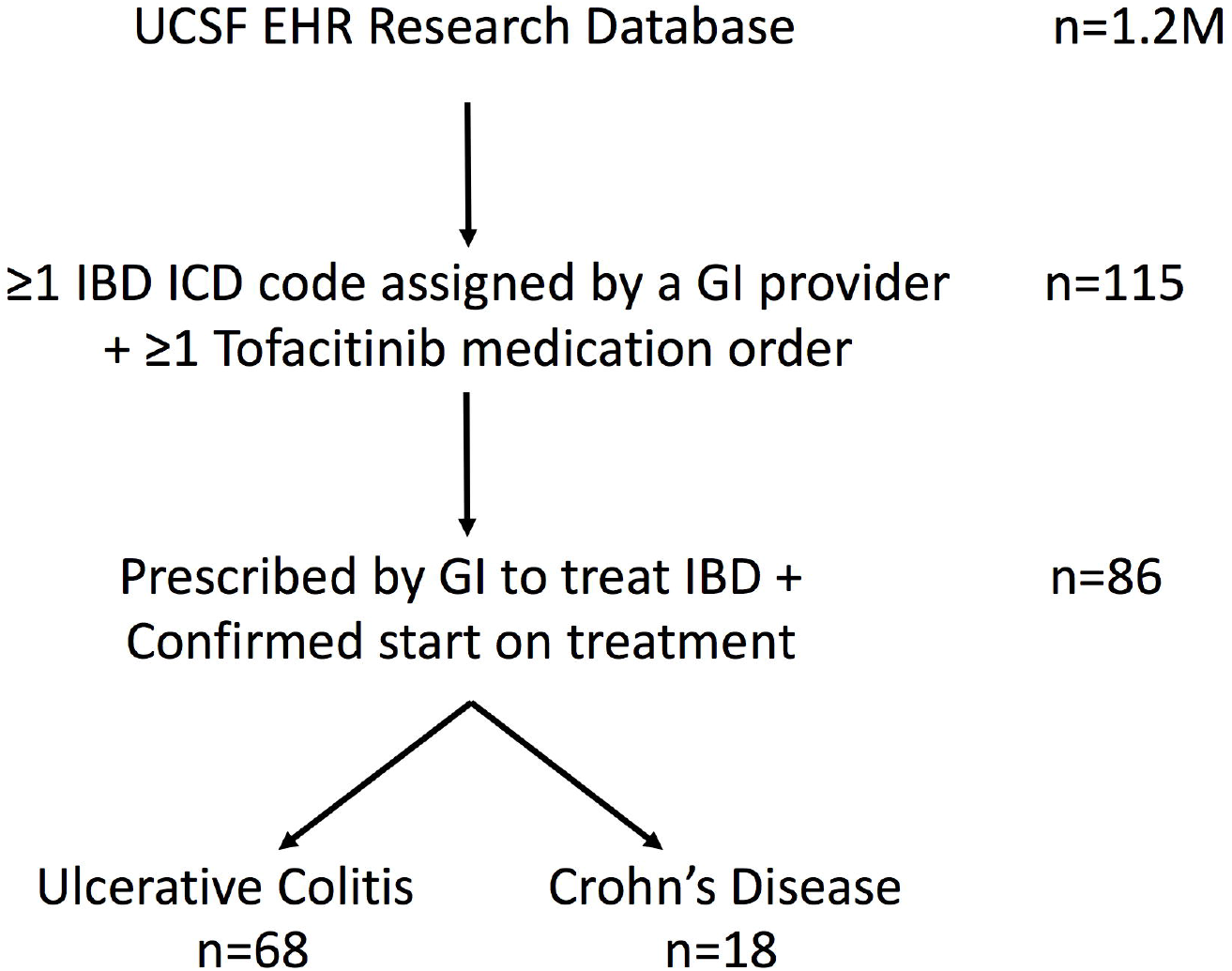
Cohort Selection Schematic.

### Comparison to RCT endpoints

We abstracted RCT endpoint definitions and rates from Sandborn et al., 2017^2^ and Panés et al., 2017^3^ (Supplemental Methods). Because both trials reported endpoint rates among those with a favorable response to treatment induction, we recalculated the overall maintenance endpoint rate as the product of the induction and maintenance response rates. All endpoints were calculated from the active treatment arm without attenuation by the placebo rate.

### Statistics and computing

We performed point estimation and hypothesis testing by calculating Wald test statistics with pooled standard errors following “Rubin’s rules”.^14^ We estimated the *time to treatment discontinuation* survival distributions using the product-limit estimator. No competing events (e.g. mortality) were observed. Treatment discontinuation due to loss of insurance coverage, as well as relocation or other lost-to-follow-up events were rare and were treated as non-informatively censored events.

We performed statistical computing in the *R* statistical computing environment (3·6·0) using the following packages: *pacman, data*.*table, survival, survminer, readxl, tidyr, scales, binom, ggplot2, lubridate, randomForest, RMarkdown, mice, Hmisc*, and *magrittr* (Supplemental References). The statistical code was independently reviewed by a co-author (BSG). Synthetic data and analysis files were version-controlled using *Docker*.

## Results

### Cohort Identification

We queried an EHR structured database system comprising ∼1·2 million patient records to identify adult patients with an IBD diagnosis assigned by a gastroenterologist and a medication order for Tofacitinib. 115 potential patient records were identified. We performed manual review to confirm that 86 patients – 68 with Ulcerative Colitis (UC) and 18 with Crohn’s Disease (CD) – had initiated Tofacitinib specifically to treat IBD (Figure 1). The other 29 patients were excluded during this process for multiple reasons, including failure to start treatment due to payor denial, the decision to forgo the ordered medical treatment in favor of surgery, and treatment initiated by a non-gastroenterologist for another autoimmune condition. Non-compliance with Tofacitinib was rare (4%) in this cohort.

### Data completeness assessment

We first sampled a subset of these 86 records to quantify the completeness of routinely collected clinical data in the capture of IBD-relevant clinical indices. We identified 87% and 91% of all the UC and CD data elements (demographics, indices) as available within the EHR. Within the set of data elements needed to calculate the baseline and follow-up Mayo Score and CDAI (for UC and CD respectively), we observed 77% and 91% data completeness. We performed multiple imputation on these missing elements to enable further downstream analysis. For instance, patients deemed treatment failures on the basis of endoscopic worsening, physician global assessment, and stool frequency but missing a rectal bleeding subscore required imputation in order to calculate the full Mayo score.

### Baseline cohort comparison

The baseline demographics of the subjects under study in the RCTs and the UCSF cohort were largely similar (Table 1). Notable differences include the universal failure of TNF inhibitors in the UCSF cohort, as well as a longer duration of disease in the UC patients. Patient groups had similar baseline Mayo scores, c-reactive protein levels, and rates of corticosteroid use.

We assessed the proportion of UC patients who would have satisfied the eligibility criteria of the corresponding Phase III RCT^2^. We found that 0% of the UC patients initiated on Tofacitinib met these criteria. The reasons for this were multifactorial (Table 2) but include prior use of Vedolizumab within the previous year, use of high-dose prednisone or intravenous methylprednisolone at the time of treatment initiation, possibility of requiring surgery during the treatment period, and the lack of protocolized screening or scheduled steroid tapering in the routine clinical setting.

**Table 2:**
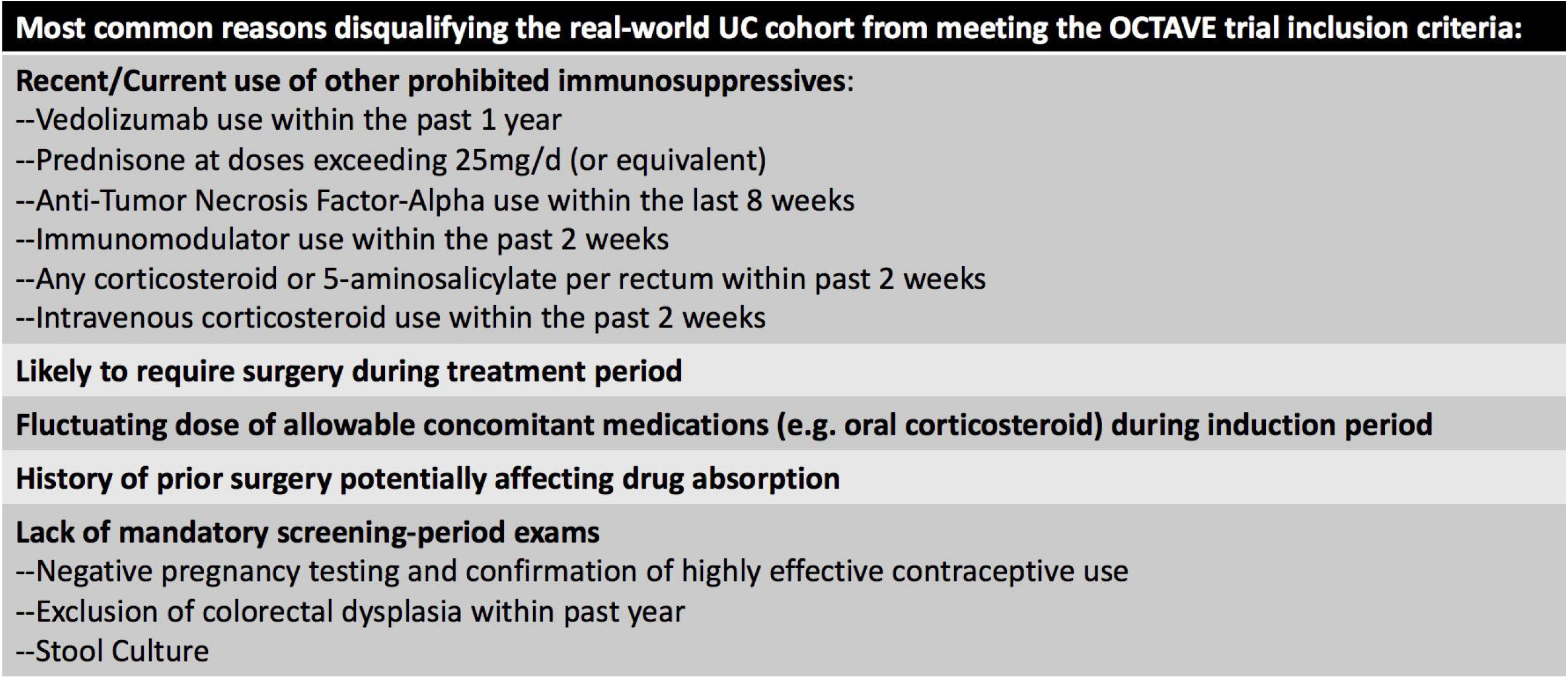
Most common reasons disqualifying the real-world ulcerative colitis cohort from meeting the OCTAVE trial eligibility criteria

We separately explored what proportion of patients met the specific RCT entry criteria defined by the Mayo score and CDAI for UC and CD respectively. 93% (73-98) of the UC patients had an eligible baseline Mayo score, whereas 50% (19-82) of the CD patients had a baseline CDAI within the eligible range of the corresponding RCT.

### Efficacy vs Effectiveness

Time to treatment discontinuation analysis on the full cohort revealed nearly identical survival distributions irrespective of IBD disease subtype (Figure 2). The overall probability of incident users maintaining long-term treatment on Tofacitinib was 68% (58-80%). All failure events occurred within the first seven months; among continued responders by month six, the probability of sustained long-term response was 94%. Of note, the first use of the Tofacitinib occurred in 2013, and the longest duration of effectiveness data relevant to treatment maintenance was 3·7 years.

**Figure 2:**
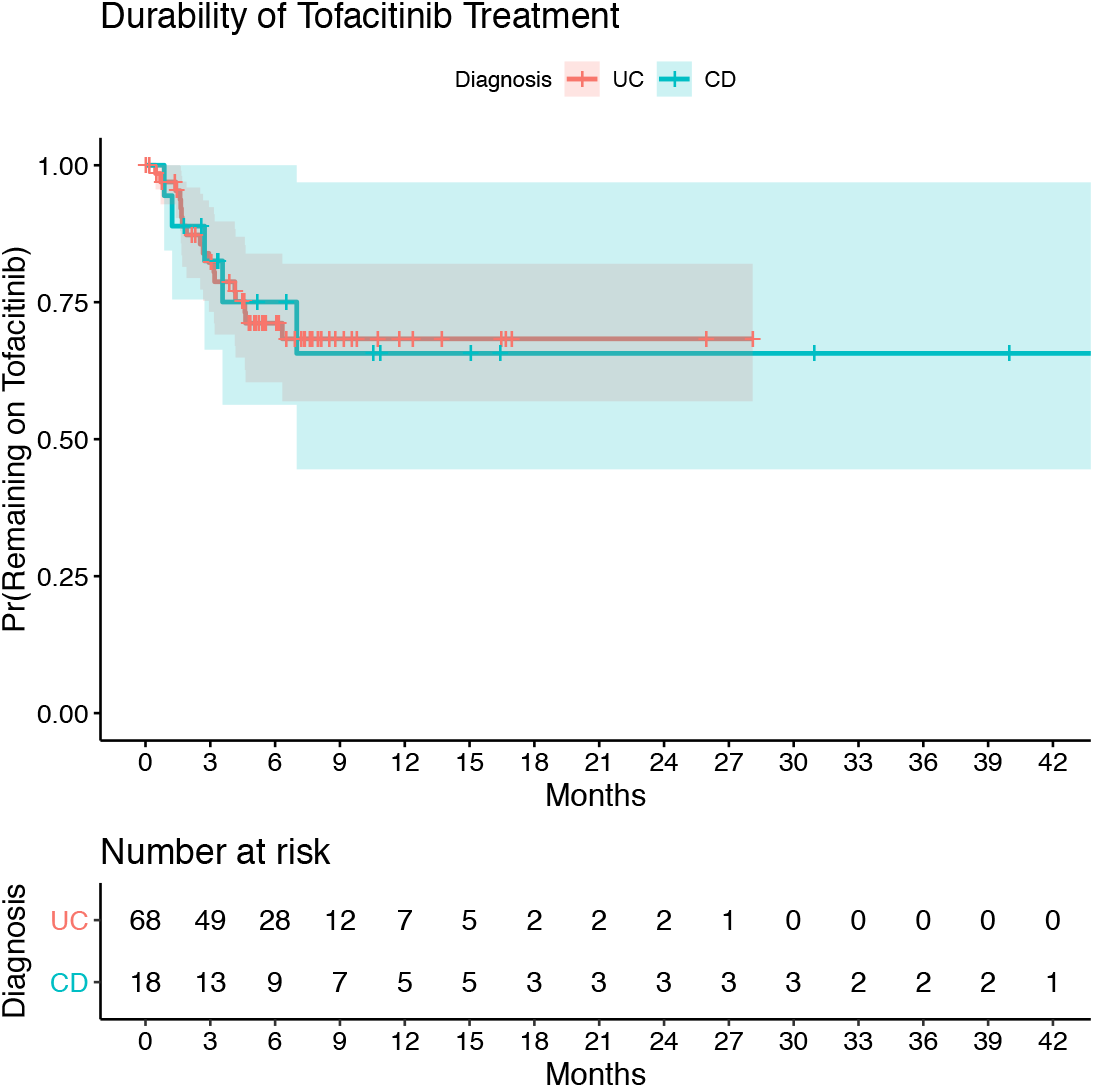
Time to Tofacitinib Discontinuation. Shaded regions correspond to the 95% confidence interval.

22% of all subjects participating in the induction phase of the UC RCT^2^ met the primary maintenance endpoint of week 52 clinical remission. We observed a similar rate (16%) in the corresponding UCSF cohort (6-37%, p-value 0·5). Similarly, the rate of achieving the primary endpoint in the CD RCT^3^ (34%) was essentially the same as the point estimate of the real-world cohort (38%, p-value 0·8).

However, the rates of meeting these regulatory endpoints were substantially different from the week 52 and week 26 survival probabilities for UC and CD respectively (p-values 3e-14 and 9e-5). This difference from the empiric response rate remained even when compared to the least stringent secondary endpoint for the UC RCT (week 52 clinical response), met by 33% of all induction participants (p-value 0·05).

## Discussion

RWD has been receiving growing interest from a variety of parties in recent years. The European Medicines Agency^15^ and the US Food and Drug Administration^7^ have been formalizing regulatory pathways to evaluate RWD in support of new drug indications and post-marketing surveillance. RWD is also being used at earlier stages of clinical development by biopharmaceuticals in order to improve drug development and clinical trial design. Healthcare payors are beginning to use RWD to support outcomes-based pricing contracts. Providers and patients are increasingly interested in understanding how RWD can enable personalized medicine in clinical practice.

Although these developments have been promising, multiple barriers have precluded the otherwise widespread adoption of RWD to improve healthcare. Missingness is ubiquitous in clinical data and especially so in the EHR, but is typically either deemphasized or handled using ad hoc and biased methods.^16^ Studies of large real-world clinical datasets typically depend on administratively coded structured data which are prone to mis-annotation.^17^ The use of registered protocols, published analysis plans, independent code review, and the public release of reproducible analysis documents are uncommon in observational studies, and have generally contributed to their decreased credibility in comparison to their better-funded, controlled study counterparts. Lastly, real-world studies have prioritized pragmatic endpoints but without sufficient attention to careful benchmarking against controlled trials using clinical instruments, further contributing to the credibility gap. Overall, there is a lack of widely-adopted standards for the proper handling of RWD to robustly support the generation of new clinical knowledge.

This study attempts to address these shortcomings and critically assess the potential of RWD to inform both research and practice via the use-case of IBD. To curtail the potential bias from unblinded record review, we published a detailed protocol and statistical analysis plan on an open platform in advance of this work. In addition to capturing the pragmatic endpoint of *time to treatment discontinuation*, we explicitly prioritized the benchmarking of our data to the corresponding RCTs and took special efforts to capture the very same clinical instruments and endpoints assessed by regulatory bodies. We critically assessed the missingness of our data and addressed it using principled methods. We performed an independent code review and have prepared a synthetic data set and dynamic analysis documents to maximize both the reproducibility of our results and the reusability of our code in other health systems.

The results of our study have important implications for both the future of real-world evidence studies and specifically the treatment of IBD. We show that routinely collected clinical data from the EHR is sufficiently rich to enable the measurement of the primary clinical instruments used in IBD RCTs with limited missingness. We also demonstrate that point-estimates of clinical effectiveness derived from RWD align well with those suggested by RCTs when using the same clinical instruments. These results support the potential for robust inference from RWD.

However, our data also speak to the presence of a significant efficacy-effectiveness gap between treatment durability in practice and regulatory endpoints measured in clinical trials. More than the regulatory endpoint, these pragmatic endpoints may provide the best answer to patients who ask: “How likely is this drug to work for me?” (Answer: “Over half of patients like you will respond to treatment long-term. It can take six months on treatment to know if you fall in this category”). Similarly, this pragmatic endpoint may be most relevant to payors increasingly keen on paying for value and anticipating the long-term costs of care.

Additionally, our study speaks to a number of implications on current trial design in IBD and their application to practice. We found that only half of the CD patients at our center mounted a CDAI score within range of the corresponding RCT. The CDAI score is known to correlate poorly with objective markers of mucosal inflammation,^18,19^ and these results overall suggest the need for improved clinical instruments for CD.^20^ We also found that 100% of UC patients at our center were “functionally disqualified” from the corresponding RCTs, consistent with prior work.^5^ On our review most of the reasons for this were related to the careful control of alternative explanations to any differences in trial endpoints seen between treatment arms. These observations reflect the fundamental trade-off between internal and external validity, and specifically the importance of integrating controlled- and real-world studies to obtain the highest quality clinical evidence.

Perhaps the most intriguing result of this study is the finding of identical survival distributions for the real-world use of Tofacitinib in CD and UC. Given the positive trial results of Tofacitinib for UC and two negative trial results in CD, we expected to see a divergence of survival distributions and a clear signal that the drug does not work in CD. This finding to the contrary suggests two important corollaries: 1) the CDAI may not be the optimal instrument to measure Crohn’s Disease activity, and 2) that the current taxonomy of IBD (UC vs CD) may not be well rooted in molecular pathogenesis. Indeed, nearly all pharmacological treatments between CD and UC are shared. Our results are consistent with recent post-hoc analyses of the CD clinical trials suggesting a signal for efficacy if analyzed using more objective endpoints,^21^ as well as the continued investigation of other JAK inhibitors for CD. These data underscore the potential of real-world studies to harness “the wisdom of the masses,” to identify robust efficacy signals that may support label expansion, and to expand options for the truly refractory patient facing limited alternatives.

We acknowledge several limitations to this study. First, our sample size is still small. This is due to the relatively recent approval of the drug for UC and its lack of EMA or FDA approval for CD. Although small samples decrease the power to detect differences, the signals seen here were strong enough to reveal many important effects, including survival distributions that are highly consistent with prior reports.^22^ Of course, patient cohorts will continue to expand as this drug continues to be used, and real-world data can keep growing.

Second, although we attempted to comprehensively characterize possible biases and place our findings in context (Table 3), we cannot exclude the possibility of residual bias. Third, the validity of imputation depends on the missing at random (MAR) assumption. We would argue that this assumption is plausible: the most common reasons for missingness were clear indications of treatment failure (e.g. no rationale to pursue endoscopy), there was complete measurement of the survival outcome, and the list of auxiliary variables was extensive. Nevertheless, it remains fundamentally untestable. Lastly, our chart review process – albeit guided by a detailed pre-published protocol – may not be entirely objective nor scalable.

**Table 3:** Qualitative Analysis of Bias

|  | Description | Effect on efficacy/effectiveness | Potential Solutions |
| --- | --- | --- | --- |
| **Confounding Bias (e.g. Bias by Indication)** |  |  |  |
| Influences on decision to treat based on likelihood of response | Many real-world patients initiated Tofacitinib following hospitalization, surgical consultation and intravenous steroids (all RCT exclusion criteria). | Possible decreased real-world effectiveness | Model-based approaches (e.g. propensity-score matching, inverse probability of treatment weighting) |
| **Selection Bias** |  |  |  |
| Restrictive RCT eligibility criteria (See Table 2) | Multiple exclusion criteria may limit generalizability to ordinary populations | Unclear effect on treatment efficacy measured by RCTs | Further studies of real-world cohorts |
| Tertiary care referral center population, off-label use in refractory patients | Greater prevalence of treatment-resistant patients | Possible decreased real-world effectiveness | Further studies of real-world cohorts |
| **Measurement Bias** |  |  |  |
| ‘Laxity’ in real-world use patterns compared to protocol-driven endpoints | Lack of steroid-de-escalation protocols and refractory patients with limited alternatives may tolerate a suboptimal treatment response for a longer period of time. | Increased real-world effectiveness measured by time to treatment discontinuation | Trend real-world response rates over time |
| Intention to treat analysis with non-responder imputation | Subjects non-adherent to the active arm and/or follow-up protocols are analyzed as treatment non-responders | Decreased treatment effect measured by RCTs | Re-analysis of clinical trial data with use of alternative imputation methods |
| Use of 6 month windows for covariate capture | Baseline and follow-up covariates were captured by protocol to accommodate real-world practice | Unclear effect on real-world effectiveness | Model-based approaches to more accurately estimate values at fixed time points |
| Unblinded chart review | Chart reviewer not blinded to outcome | Unclear effects on real-world effectiveness | Use of multiple blinded reviewers (assessing unrelated components) with measurement of interrater reliability |

In summary, our study suggests that RWD can be a robust source of valuable clinical knowledge that complements evidence from controlled trials. Tofacitinib appears to work as well as has been suggested by clinical trials when using the same clinical instruments, but has greater real-world durability. Despite negative trial results, Tofacitinib and JAK inhibitors more generally may be valuable for the treatment of CD. Lastly, because clinical trials functionally exclude many patients, real-world studies are indispensable to ensure the generalizability of RCT findings and add to the best evidence for clinical care.

## Data Availability

The analytic code in the form of a R markdown file as well as the accompanying data set needed to reproduce the analysis in this work are available in a Docker container and will be released for public use on https://datadryad.org at the time of publication to all investigators without restriction (https://doi.org/10.7272/Q6PZ5715). These individual participant data were de-identified to comply with the US Department of Health and Human Services 'Safe Harbor' guidance and applicable laws and regulations concerning privacy and/or security of personal information. The data dictionary is documented within the study protocol section of Supplemental Content.

## Author Contributions

VAR and AJB conceived the project. VAR designed the chart review protocol, performed chart extraction, conducted statistical analyses, and drafted this manuscript. BSG performed code review and critically edited this manuscript. AJB supervised the project and critically edited this manuscript.

## Acknowledgements

The authors thank the UCSF Academic Research Services and Clinical Data Research Consultation services for clinical informatics support. The authors would like to acknowledge Dana Ludwig for his help in deidentifying and interpreting the UCSF EHR.

## Data Sharing Plan

The analytic code in the form of a R markdown file as well as the accompanying data set needed to reproduce the analysis in this work are available in a *Docker* container and will be released for public use on https://datadryad.org at the time of publication to all investigators without restriction (https://doi.org/10.7272/Q6PZ5715). These individual participant data were de-identified to comply with the US Department of Health and Human Services ‘Safe Harbor’ guidance and applicable laws and regulations concerning privacy and/or security of personal information. The data dictionary is documented within the study protocol section of Supplemental Content.

